# ENRICHing Medical Imaging Training Sets Enables More Efficient Machine Learning

**DOI:** 10.1101/2021.05.22.21257645

**Authors:** Erin Chinn, Rohit Arora, Ramy Arnaout, Rima Arnaout

## Abstract

**Objective:** Deep learning (DL) has been applied in proofs of concept across biomedical imaging, including across modalities and medical specialties^1–17^. Labeled data is critical to training and testing DL models, but human expert labelers are limited. In addition, DL traditionally requires copious training data, which is computationally expensive to process and iterate over. Consequently, it is useful to prioritize using those images that are most likely to improve a model’s performance, a practice known as instance selection. The challenge is determining how best to prioritize. It is natural to prefer straightforward, robust, quantitative metrics as the basis for prioritization for instance selection. However, in current practice such metrics are not tailored to, and almost never used for, image datasets.

**Methods:** To address this problem, we introduce ENRICH—<u>E</u>liminate <u>N</u>oise and <u>R</u>edundancy for Imaging <u>Ch</u>allenges—a customizable method that prioritizes images based on how much diversity each image adds to the training set.

**Results:** First, we show that medical datasets are special in that in general each image adds less diversity than in non-medical datasets. Next, we demonstrate that ENRICH achieves nearly maximal performance on classification and segmentation tasks on several medical image datasets using only a fraction of the available images and outperforms random image selection, the negative control. Finally, we show that ENRICH can also be used to identify errors and outliers in imaging datasets.

**Conclusion:** ENRICH is a simple, computationally efficient method for prioritizing images for expert labeling and use in DL.

## Background and Significance

Deep learning (DL) models can classify images by disease or by the structure(s) they contain. They can also segment, track, and measure substructures within images^1–17^. DL thus has great promise for helping meet the overwhelming need for accurate, reliable, and scalable image interpretation that currently exists in medicine due to a near-universal shortage of trained human experts^5,6,18–21^. However, the data-hungry nature of DL model training threatens to hamper its effective use for medical imaging. First, the large amount of data required creates a costly and time-consuming labeling bottleneck for clinical experts^3^. (This is in contrast to labeling in non-medical fields, which usually focuses on everyday objects and therefore can be performed more quickly and inexpensively by laypeople via crowdsourcing^22^.) Various unsupervised methods can help mitigate the labeling burden but cannot eliminate it, since experts are still needed to label test datasets in order to benchmark performance on high-stakes medical tasks. Second, large amounts of training data increase training times, slowing iteration during model development, requiring out-of-reach computational resources, or both. While early proofs of concept have used relatively small imaging datasets, the skyrocketing volume of medical image data promises to magnify these challenges.^23,24^

It has long been recognized that prioritizing the labeling of data that most benefit model performance, a practice known as *instance selection*, as opposed to random data, can be helpful for machine learning^25,26^. Instance selection methods generally balance some measure of the representativeness of a datapoint (instance) with some measure of how much that instance will add to the diversity of the resulting training set^25^. In contrast to *active learning*,^27,28^ which requires iterative training of DL models to identify additional instances to include, instance selection algorithms are used once up front. Most work on instance selection has focused on non-imaging data and preceded recent developments in DL. In addition, instance selection methods described to date often require prior knowledge of the instance label and so would not reduce the data labeling burden. Therefore, how best to use instance selection for images, and how best to curate large image datasets in a label-free approach, are open questions.

Medical images differ from images of everyday objects in ways that we hypothesized could be leveraged for a new instance selection approach. Unlike images of everyday objects, which typically exhibit multiple lighting conditions and are captured at a range of distances, angles, and contexts, medical images are often more uniform in these respects, a result of standardization of imaging protocols for patient care. Images from a particular medical domain often have similar subject matter (e.g., the heart in cardiology, the retina in ophthalmology), pose (standard views), background (black), noise, lighting, and color. In the case of computed tomography (CT), magnetic resonance imaging (MRI), ultrasound (US), and other common imaging modalities, image frames are often captured consecutively, resulting in similarity among images. For these reasons, we hypothesize that standardization in medical imaging creates greater redundancy in medical training data than in commonly used non-medical datasets. We therefore propose that simply prioritizing non-redundant images is an efficient means of instance selection for DL in medical imaging.

As a test of this understanding, here we present a method called ENRICH: Eliminating Noise and Redundancy for Imaging Challenges. It consists of two main steps. First, a similarity metric is calculated for all pairs of images in a given dataset, forming a matrix of pairwise-similarity values. Second, an algorithm operates on the matrix to identify those images that are least similar to images in an existing seed training set and thereby hypothetically most informative. The result is a meaningful decrease in the redundancy and size of the resulting training set without requiring up-front labeling. We demonstrate proof of concept on classification and segmentation tasks on two large, well characterized medical datasets: ECHO-F^3^, which consists of fetal echocardiograms, and OCT^29^, which consists of adult retinal optical coherence tomography images. We also demonstrate the special nature of medical image datasets, showing differences in their pairwise similarities compared to STL10, a standard non-medical image dataset used for various DL applications^30^.

## Materials and Methods

### Datasets, tasks, and benchmarks

We searched for available datasets meeting the following criteria: minimum image size of 80×80 pixels; minimum image number 10,000 (classification) or 1,000 (segmentation); not trivially simple (e.g., MNIST); multiple labeled classes available; representing both non-medical images as well as medical images of different anatomic structures, different imaging modalities, and different data structure (still images vs videos). Training and test sets are described below and in **Table 1**.

**Table 1.** Overall training and testing datasets.

| Dataset | Class | Training |  |  | Testing |  |  |
| --- | --- | --- | --- | --- | --- | --- | --- |
|  |  | Images | Clips | Patients | Images | Clips | Patients |
| **ECHO-F** | 3VV | 7159 | 837 | 500 | 890 | 96 | 51 |
|  | A4C | 20378 | 1495 | 652 | 3518 | 198 | 80 |
|  | NT | 25082 | 2849 | 281 | 4365 | 764 | 51 |
|  | *Total\** | 52619 | 4687 | 777 | 8773 | 976 | 89 |
| **ECHO-F-SEG** | A4C | 1248 | 299 | 186 | 173 | 48 | 20 |
| **OCT** | NL | 23468 | -- | 3193 | 3015 | -- | 433 |
|  | CNV | 22696 | -- | 653 | 14623 | -- | 267 |
|  | DME | 6994 | -- | 601 | 4550 | -- | 166 |
|  | DRU | 4144 | -- | 574 | 4720 | -- | 142 |
|  | *Total\** | 57302 | -- | 4221 | 26908 | -- | 1254 |
| **STL10** | AIR | 6059 | -- | -- | 800 | -- | -- |
|  | TRUCK | 4117 | -- | -- | 800 | -- | -- |
|  | SHIP | 6600 | -- | -- | 800 | -- | -- |
|  | *Total* | 16776 | -- | -- | 2400 | -- | -- |

\* some clips and patients may contain more than one class of image
3VV, 3-vessel view; A4C, apical 4-chamber; NT, non-target; NL, normal; CNV, choroidal neovascularization; DME, diabetic macular edema; DRU, drusen; AIR, airplane.

#### ECHO-F

ECHO-F consists of fetal echocardiogram images^3^. The binary classification task included the fetal axial 4-chamber (A4C) view and the non-target (NT) view. The multi-class task included the A4C and NT views as well as the fetal 3-vessel view (3VV). In ultrasound, one or more clips are acquired per patient; each clip consists of one to several hundred consecutive image frames. Training and test sets were divided by patient identifier (ID) and were disjoint from each other.

#### ECHO-F-SEG

ECHO-F-SEG consists of a subset of A4C images from ECHO-F. ECHO-F-SEG was used for multi-class segmentation with 5 classes: left ventricle, right ventricle, left atrium, right atrium, and background. Notably, ECHO-F-SEG had already been curated informally, in that only certain frames from each video clip were labeled.

#### OCT

OCT consists of adult retinal optical coherence tomography images^29^. Binary classification was between normal retina (NL) and choroidal neovascularization (CNV). The multi-class task included NL and CNV as well as drusen (DRU) and diabetic macular edema (DME). The train/test split of the dataset was adjusted from the authors’ original description: instead of 250 images per lesion in the test set, we created disjoint train/test sets as we did for ECHO-F, split by patient ID, and increased the total size of the binary test set from 500 images to 17,638 images and the multi-class test set from 1,000 images to 26,908 images to make the tests more difficult.

#### STL10

STL10 consists of images of animals and vehicles^30^. The binary classification task included images of airplanes (AIR) and trucks (TRUCK). The multi-class task included AIR, TRUCK, and ships/boats (SHIP).

### Image processing

Grayscale conversion was done using Python3’s OpenCV package. Image resizing was done using Python3’s Scikit-Image package.

#### ECHO-F

ECHO-F images were originally 300×400 pixel greyscale. For autoencoder input, the original images were cropped to square and resized to 64×64. For classification-model training and testing, images were cropped and resized to 80×80. For segmentation-model training and testing, original images were cropped to 272×272.

#### OCT

OCT images were originally grayscale and varied in pixel dimension. This dataset was standardized to correct region-of-interest misalignment and remove white-edge artifacts (discovered, in fact, by ENRICH). First, white sections at image edges were removed, then images underwent a cropping and resizing as above.

#### STL10

STL10 images were originally 96×96 and converted to grayscale to for ease of comparison with the medical image datasets and resized as above.

### Embeddings

The bottleneck layer of a disentangled variational autoencoder (β-VAE) was used to compress each image into a 128-element vector embedding. The β-VAE used was based on the architecture as described previously except for the dimension of the embedding^31^. The β-VAE was trained on a subset of 5,000 images from the entire ECHO-F training dataset as previously described^3^, using combined loss (reconstruction loss and Kullback-Leibler divergence) and standard stopping conditions. Autoencoders trained with different images produced comparable embeddings (not shown).

### Pairwise image similarities

For each dataset, a matrix of pairwise image similarities was calculated. The similarity between two image embeddings was defined as the cosine similarity (the complement of the cosine distance) between each embedding, resulting in pairwise similarities ranging from 0 for highly dissimilar images to 1 for identical images.

### Ranking algorithm

For each deep-learning task, an initial subset of images was chosen at random. For each image *i* in the remaining dataset, the maximum similarity to each image *j* in the training set, max_*j*_(*z*_*ij*_), was read from the similarity matrix, and the image *i* with the smallest max_*j*_(*z*_*ij*_) was added to the training set (i.e., argmin_*i*_(max_*j*_(*z*_*ij*_))). This step was iterated to grow the training set, and the quality of the training set was assessed at specific sizes by training a model and measuring its performance (below). The ranking algorithm was blind to class label. For statistical confidence, the ranking algorithm, including choice of the initial subset, was repeated 3 times for each task (“biological” replicates), and 10 models were trained on each resulting training set (technical replicates). Training subsets are summarized in **Tables 2-4**.

**Table 2.** Select training subsets, ECHO-F.

| **ECHO-F binary** |  |  |  |  |  |  |  |  |  |  |  |
| --- | --- | --- | --- | --- | --- | --- | --- | --- | --- | --- | --- |
| **No. Images** | **1000\*** | **3000** | **5000** | **10000** | **15000** | **20000** | **25000** | **30000** | **35000** | **40000** | **45460** |
| Pct. Images | 2 | 7 | 11 | 22 | 33 | 44 | 55 | 66 | 77 | 88 | 100 |
| No. Images A4C (avg±sd) | 444±7 | 1019±7 | 1533±11 | 2588±8 | 4352±22 | 6531±13 | 9108±160 | 11833±18 | 15260±8 | 20260±8 | 20378±0 |
| No. Images NT (avg±sd) | 556±7 | 1981±7 | 3467±11 | 7412±8 | 10648±22 | 13469±13 | 15891±160 | 18167±18 | 19739±8 | 19739±8 | 25082±0 |
| No. Clips A4C (avg±sd) | 347±16 | 882±21 | 1319±1 | 1480±2 | 1493±1 | 1495±0 | 1495±0 | 1495±0 | 1495±0 | 1495±0 | 1495±0 |
| No. Clips NT (avg±sd) | 424±13 | 1346±11 | 2139±9 | 2741±3 | 2794±2 | 2810±1 | 2817±3 | 2826±1 | 2833±1 | 2833±1 | 2849±0 |
| No. Clips all classes (avg±sd) | 770±9 | 2186±12 | 3381±7 | 4109±3 | 4171±2 | 4190±1 | 4197±3 | 4206±1 | 4213±1 | 4213±1 | 4229±0 |
| No. Patients A4C (avg±sd) | 266±14 | 502±8 | 630±1 | 652±0 | 652±0 | 652±0 | 652±0 | 652±0 | 652±0 | 652±0 | 652±0 |
| No. Patients NT (avg±sd) | 104±1 | 222±6 | 269±1 | 281±1 | 281±0 | 281±0 | 281±0 | 281±0 | 281±0 | 281±0 | 281±0 |
| No. Patients all classes (avg±sd) | 337±10 | 592±9 | 697±2 | 713±0 | 713±0 | 713±0 | 713±0 | 713±0 | 713±0 | 713±0 | 713±0 |
| **ECHO-F multi-class** |  |  |  |  |  |  |  |  |  |  |  |
| **No. Images** | **1000\*** | **3000** | **5000** | **10000** | **15000** | **20000** | **25000** | **30000** | **35000** | **40000** | **52619** |
| Pct. Images | 2 | 6 | 10 | 19 | 29 | 38 | 48 | 57 | 67 | 76 | 100 |
| No. Images 3VV (avg±sd) | 139±2 | 461±17 | 742±7 | 1174±2 | 1728±16 | 2391±6 | 3899±8 | 3899±8 | 4788±1 | 6937±13 | 7159±0 |
| No. Images A4C (avg±sd) | 381±6 | 847±17 | 1312±4 | 2306±8 | 3666±13 | 5976±28 | 7388±5 | 10593±33 | 12993±25 | 13244±28 | 20378±0 |
| No. Images NT (avg±sd) | 480±6 | 1692±7 | 2946±4 | 6520±9 | 9606±14 | 11633±25 | 13713±9 | 15508±26 | 17219±24 | 19819±18 | 25082±0 |
| No. Clips 3VV (avg±sd) | 118±6 | 423±9 | 676±3 | 832±1 | 836±0 | 837±1 | 837±0 | 837±0 | 837±0 | 837±0 | 837±0 |
| No. Clips A4C (avg±sd) | 307±3 | 741±15 | 1155±2 | 1476±2 | 1489±2 | 1494±1 | 1495±1 | 1495±1 | 1495±0 | 1495±0 | 1495±0 |
| No. Clips NT (avg±sd) | 378±5 | 1181±11 | 1907±5 | 2697±6 | 2783±2 | 2798±2 | 2809±1 | 2817±1 | 2820±1 | 2830±0 | 2849±0 |
| No. Clips all classes (avg±sd) | 792±8 | 2242±10 | 3494±5 | 4523±6 | 4618±1 | 4635±0 | 4647±1 | 4655±1 | 4658±0 | 4668±0 | 4687±0 |
| No. Patients 3VV (avg±sd) | 103±8 | 313±5 | 444±6 | 500±0 | 500±0 | 500±0 | 500±0 | 500±0 | 500±0 | 500±0 | 500±0 |
| No. Patients A4C (avg±sd) | 246±5 | 447±10 | 588±3 | 652±1 | 652±0 | 652±0 | 652±0 | 652±0 | 652±0 | 652±0 | 652±0 |
| No. Patients NT (avg±sd) | 98±9 | 206±2 | 251±1 | 281±0 | 281±0 | 281±0 | 281±0 | 281±0 | 281±0 | 281±0 | 281±0 |
| No. Patients all classes (avg±sd) | 371±7 | 629±5 | 746±3 | 777±0 | 777±0 | 777±0 | 777±0 | 777±0 | 777±0 | 777±0 | 777±0 |
| **ECHO-F-SEG** |  |  |  |  |  |  |  |  |  |  |  |
| **No. Images** | **200\*** | **300** | **400** | **600** | **800** | **1000** | **1050** | **1100** | **1150** | **1200** | **1248** |
| Pct. Images | 16 | 24 | 32 | 48 | 64 | 80 | 84 | 88 | 92 | 96 | 100 |
| No. Images A4C (avg±sd) | 200±0 | 300±0 | 400±0 | 600±0 | 800±0 | 1000±0 | 1050±0 | 1100±0 | 1150±0 | 1200±0 | 1248±0 |
| No. Clips A4C (avg±sd) | 121±4 | 156±8 | 180±7 | 224±7 | 260±5 | 281±2 | 285±3 | 289±2 | 294±1 | 298±1 | 299±0 |
| No. Patients A4C (avg±sd) | 100±3 | 121±7 | 130±6 | 151±5 | 167±1 | 178±2 | 179±2 | 181±2 | 184±1 | 185±0 | 186±0 |

3VV, 3-vessel view; A4C, axial 4-chamber; NT, non-target
\* initial “seed” subset

**Table 3.**
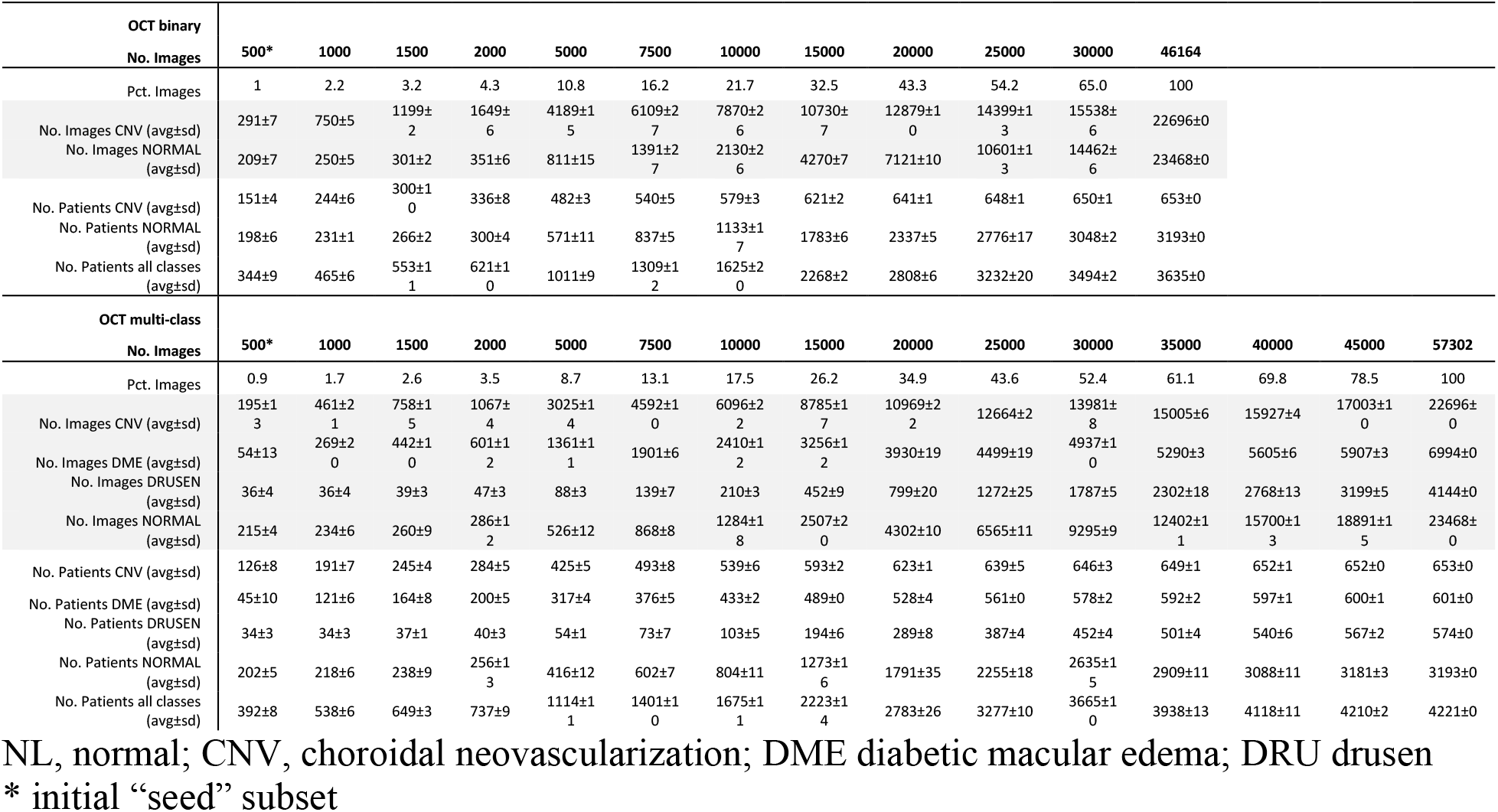
Select training subsets, OCT.

**Table 4.** Select training subsets, STL10.

| STL10 binary |  |  |  |  |  |  |  |  |  |  |
| --- | --- | --- | --- | --- | --- | --- | --- | --- | --- | --- |
| No. Images | 500* | 1000 | 2000 | 3000 | 5000 | 7000 | 9000 | 10176 |  |  |
| Pct. Images | 4.9 | 9.8 | 19.7 | 29.5 | 49.1 | 68.8 | 88.4 | 100 |  |  |
| No. Images AIRPLANE (avg±sd) | 299±6 | 407±5 | 632±7 | 946±4 | 1887±3 | 3457±1 | 5362±1 | 6059±0 |  |  |
| No. Images TRUCK (avg±sd) | 201±6 | 593±5 | 1368±7 | 2054±4 | 3113±3 | 3543±1 | 3638±1 | 4117±0 |  |  |
| STL10 multi-class |  |  |  |  |  |  |  |  |  |  |
| No. Images | 500* | 1000 | 2000 | 3000 | 5000 | 7000 | 9000 | 11000 | 13000 | 16776 |
| Pct. Images | 3 | 6 | 11.9 | 17.9 | 29.8 | 41.7 | 53.6 | 65.6 | 77.5 | 100 |
| No. Images AIRPLANE (avg±sd) | 183±14 | 260±15 | 438±7 | 623±11 | 1087±16 | 1706±8 | 2436±33 | 3323±10 | 4290±10 | 6059±0 |
| No. Images SHIP (avg±sd) | 194±7 | 329±8 | 566±21 | 857±8 | 1531±18 | 2304±14 | 3215±32 | 4160±10 | 5109±10 | 4117±0 |
| No. Images TRUCK (avg±sd) | 123±9 | 411±14 | 996±16 | 1520±3 | 2381±3 | 2990±7 | 3349±6 | 3517±4 | 3601±1 | 6600±0 |

\* initial “seed” subset

### Model training

Resnet and U-net architectures were used to train classification and segmentation models, respectively, as previously described^3^. Data augmentation was used for the segmentation task as previously described^3^ but not for the classification tasks. Experiments for each dataset used the same model parameters throughout.

### Human labeling time estimates

Human labeling time averaged across *n*=4 labelers using several different labeling platforms was 3 seconds per image for classification in ECHO-F and 5 minutes per image for ECHO-F-SEG segmentation.

### Evaluation metrics

For the binary classification tasks, model performance was assessed using the area under the receiver operator characteristic curve (AUC). For the multi-class tasks, a per class one-vs-rest AUC was averaged and compared between models. For the segmentation task, average Jaccard score of the four heart segments (left ventricle, right ventricle, left atrium, right atrium), was used, as previously described^3^. One-sided t-tests were used to compare performance across experiments.

Several datasets contained hierarchical levels of organization, image < clip < patient. Representativeness of each training subset by level as applicable was calculated as a percentage of each of these in overall training set. Class balance was calculated as the effective number^32^ of classes, specifically ^1^*D*, i.e. exponentiating the Shannon entropy of the classes. At each level, class balance was calculated from the distribution of the number of unique images/clips/patients in each class.

## Results

### Dataset diversity plots and the dataset diversity score

We developed a new way to visualize the diversity or redundancy of a dataset by plotting a cumulative histogram of the values in the similarity matrix, resulting in a *dataset diversity plot* (**Fig. 1**). In such plots, high-diversity datasets—ones in which the images are very different from each other—will trace out curves to the upper left, while high-redundancy datasets will trace out curves to the lower right. The area under the diversity curve ranges from 0 to 1: 0 for datasets that are completely redundant and 1 for datasets that are maximally diverse. Thus, we used the area under the diversity curve as a natural a *dataset diversity score*, as we introduce and illustrate in **Fig. 1a**. We created diversity plots and dataset diversity scores for each dataset in our study (**Fig. 1b**). These plots revealed significant redundancy in the medical image datasets ECHO-F and OCT (**Fig. 1b, blue and green**), and higher redundancy in these medical datasets than in the non-medical STL10 (**Fig. 1b, yellow**). To our knowledge this is the first quantitative demonstration of diversity and the first comparison of diversity between medical and non-medical image datasets.

**Figure 1.**
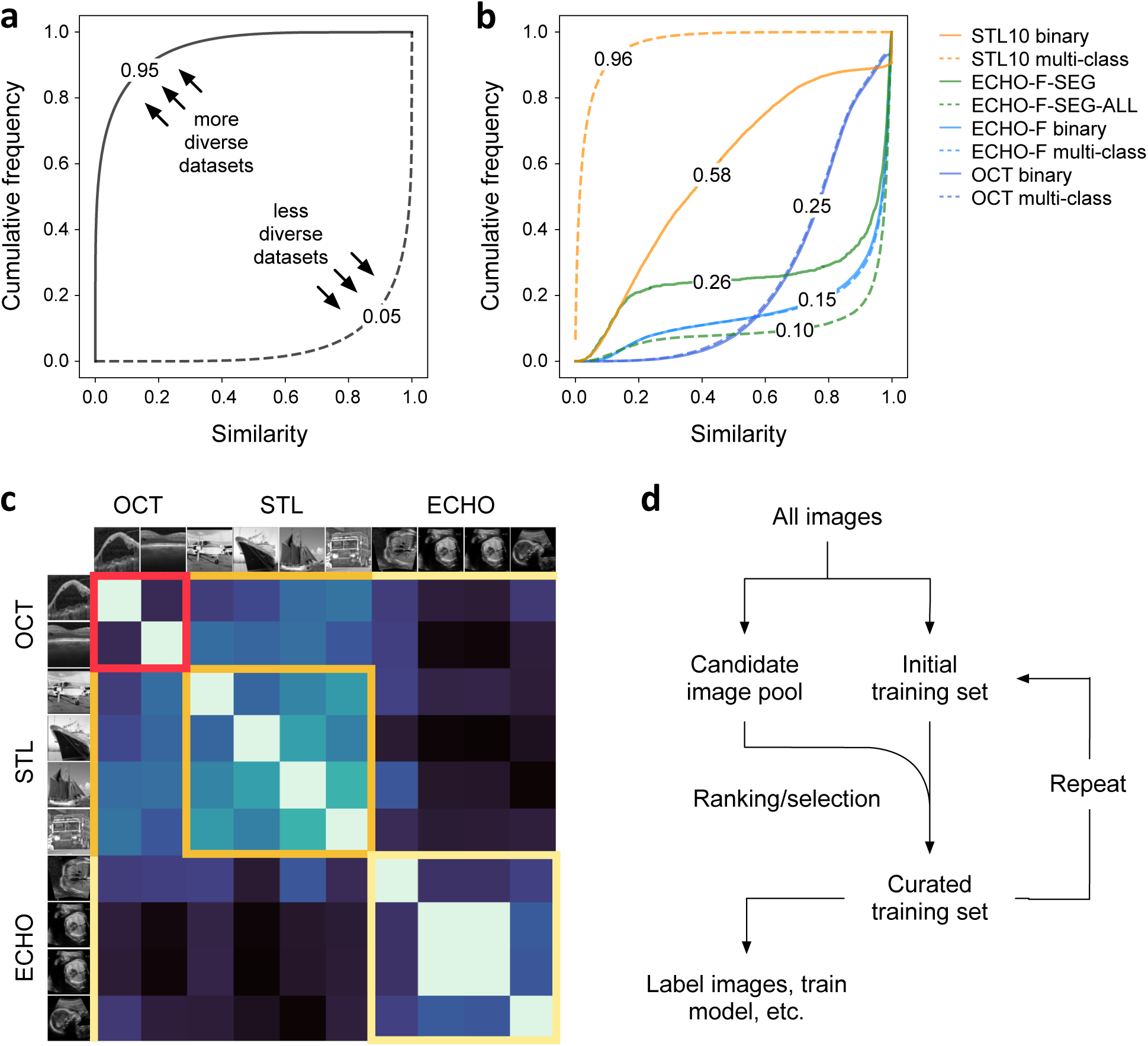
Similarity in imaging datasets and experimental approach. (a) Schematic of the dataset diversity plot, a cumulative density plot of maximum pairwise similarities. Dataset diversity scores are indicated. (b) Dataset diversity plots and scores for ECHO-F, OCT, STL10, and ECHO-F-SEG datasets. Also included are the total images available for the ECHO-F segmentation task, ECHO-F-SEG-ALL. (c) Pairwise image similarities in a handful of images drawn from OCT, STL10, and ECHO-F. Red-, orange-, and yellow-bordered squares indicate similarities within the OCT, STL, and ECHO datasets, respectively. (d) Schematic summary of ENRICH. From all available images in a dataset, an initial training set is chosen at random. The remaining images comprise a candidate pool of images from which additional images can be selected. A matrix of pairwise image similarities (Step 1 of ENRICH) is constructed. From this matrix, an algorithm is used to choose additional images to add to the initial training set; this is Step 2 of ENRICH. This process is repeated, iteratively adding images to an initial subset.

### Image redundancy in medical datasets

Our prior work suggested that medical image datasets are often quite redundant compared to non-medical datasets^3^ and that such redundancy is not confined to images from a given patient or video clip but instead is distributed across the dataset. To test this hypothesis, for each dataset we calculated the maximum pairwise similarity for each image, with a similarity measure based on the cosine similarity between β-VAE embeddings (Methods). Supporting this hypothesis, we found that the majority of ECHO-F classification images had a maximum similarity greater than 0.9: most images had at least one other image in the dataset to which they were at least 90% similar (**Fig. 1b**). The OCT dataset exhibited the same phenomenon, with roughly half of the images having a maximum pairwise similarity greater than 0.8. In contrast, most images in the non-medical STL10 dataset had a maximum similarity less than 0.4 (**Fig. 1b**).

### Comparing ENRICH-curated training subsets vs. control

ENRICH involves ranking images based on their similarity to each other and preferentially choosing the most unique—i.e., lowest similarity—images for inclusion in the training set (Methods). Labeling is not required. For each dataset, we compared the performance of models trained on ENRICHed subsets of the dataset to those trained on control subsets, i.e. subsets created by random sampling of the full dataset. We further compared the performance of both ENRICHed and control subsets of different sizes to the performance of models trained on the full training set, i.e. all available training images, as the gold standard. For each dataset and task, we recorded whether and at what size ENRICHed subsets outperformed control subsets, as well as the minimum size at which ENRICHed subsets performed indistinguishably from the gold standard (full dataset). We tested binary and multi-class classification as well as segmentation, with replicates for statistical confidence (**Fig. 2**).

**Figure 2.**
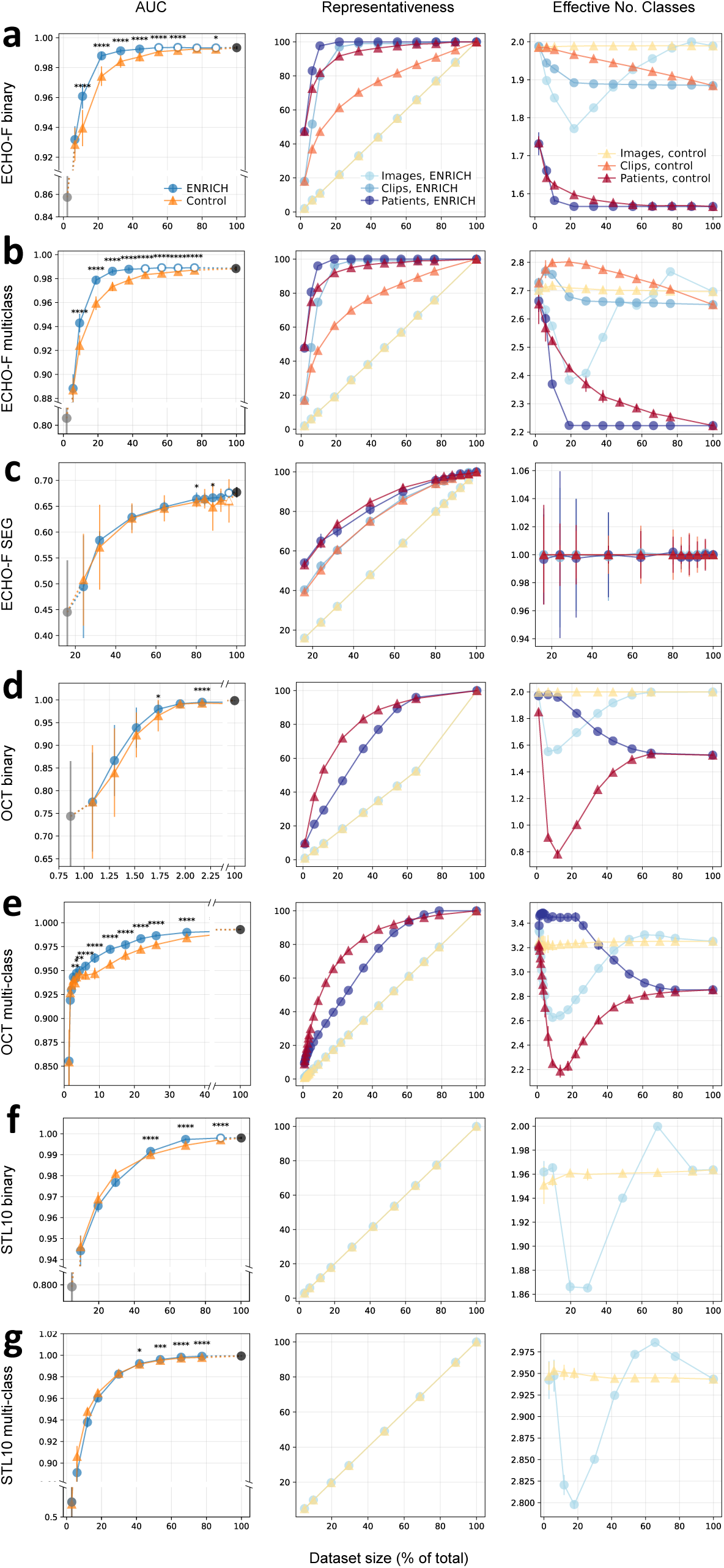
Performance of ENRICHed training datasets compared to randomly selected training datasets. (a) ECHO-F binary, (b) ECHO-F multi-class, (c) ECHO-F-SEG segmentation, (d) OCT binary, (e) OCT multi-class, (f) STL10 binary, and (g) STL10 multi-class. Each panel shows test performance on top, representativeness of images in the middle, and effective class size on the bottom. Performance testing, top: from a common initial random starting dataset (grey), additional images were added to grow increasingly larger training subsets using ENRICH (blue) vs random addition (yellow). Each datapoint represents mean AUC on the test set from 30 replicates; error bars for each datapoint show one standard deviation around the mean. Asterisks for each training data subset represent statistical differences between ENRICH and random according to the standard convention (*n*s = *p*>0.05; *= *p*≤0.05; **= *p*≤0.01; ***= *p*≤0.001; ****= *p*≤0.0001). Empty symbols are statistically indistinguishable from model performance using the full training set (100 percent of training images; black dot). Representativeness, middle: for ENRICH (cool colors) and random selection (warm colors), for each training subset, the percentage of the total training set is shown at an image (light blue, light yellow), clip (medium blue, orange), and patient (dark blue, red) levels where applicable). Effective number of classes, bottom: for ENRICH (cool colors) and random selection (warm colors), for each training subset, the effective number of classes is shown at an image (light blue, light yellow), clip (medium blue, orange), and patient (dark blue, red) levels where applicable). For representativeness and effective size as well, error bars are shown but are small, and relevant p-values are summarized in the text.

### ENRICH achieved gold-standard performance with substantially smaller training sets and outperformed unbiased selection

ENRICHed subsets achieved gold-standard performance using training subsets there were only a fraction of the size of the full training set (**Fig. 2**). Specifically, ENRICH required only 55 percent and 48 percent of available images for ECHO-F binary and multi-class tasks, respectively (**Fig. 2**). In contrast, random sampling failed to reach this benchmark at any training set size short of the full dataset. ENRICH outperformed control on even the smallest subsets, for example 11 percent of ECHO-F in the binary classification task, 9.5 percent of ECHO-F in multi-classification (p-values < 2×10^−9^; **Fig. 2a-2b**).

### ENRICH discovers dataset structure without labels

Like many medical datasets, ECHO-F has hierarchical structure, with individual video frames stratified by clip and patient. We found that ENRICH selected images that represented almost all of the available patients and video clips even at small subset sizes, significantly more so than random sampling (all p-values <1×10^−4^) (**Table 2; Fig. 2a-2b**). The largest magnitude in those differences were evident in the smallest training subsets that first showed performance gains over random selection.

### ENRICH achieves class balance without labels

Class balance was measured as the effective number^32–35^ of classes at the image, clip, and patient levels (**Fig. 2**). Effective number is a mathematically rigorous standard that takes similarities and frequencies into account. (For example, if the frequencies of the two classes in a binary classification set were highly imbalanced at 0.99 and 0.01, the effective number of classes will be close to 1, not 2, since to a good approximation—”in effect”—only the first class is represented^32^.) Using Shannon entropy (see Methods), we calculated the effective number of classes represented by the images, clips, and patients in the full training set and in each ENRICHed and control subset.

The number of images in the full training set for ECHO-F were nearly equally balanced between classes, and indeed the effective number of classes in the binary classification task is approximately two (1.99). The effective number of classes at the clip and patient levels in the full training set are 1.88 and 1.57, respectively. While random sampling saw image-level effective size stay constant and clip- and patient-level effective sizes decrease approximately linearly, ENRICHed training sets were significantly less diverse at image, clip, and image levels, most notably at the 11 and 22 percent training subsets where ENRICH first pulled ahead in binary classification performance (**Fig. 2a**; p-values all < 5×10^−4^). Similar behavior was present in the 9.5 percent subset in the multi-classification task (**Fig. 2b**; all p-values <0.001). For both tasks, imbalance favored the NT class (**Table 2**), which clinically is felt to be more diverse, since it can contain any non-heart image, while the A4C class contains only A4C images of the heart. Thus, ENRICH selectively enriched training sets for the more diverse class, resulting in better performance, as opposed to blindly maintaining class balance.

### Performance on additional datasets

Model test performance on OCT binary classification achieved a mean AUC of 0.99 (± 2.24×10^−5^) when trained on the full training dataset. ENRICH outperformed control training sets at just 2 percent of all OCT images (mean AUC 0.995 vs 0.993, p-value 9.98×10^−6^). Only 32.5 percent of the training dataset was needed to achieve gold-standard performance when training images were chosen using ENRICH vs. 41 percent for the control. The OCT dataset structure had images and patients, but not clips (**Table 1**). At a patient level, the effective number of classes is only 1.5 due to more patients in the NORMAL class than in the CNV class. For OCT, the effective number of classes patients represented in ENRICHed subsets was higher even as control subsets had a higher effective number of classes among images (**Fig. 2d**). As Table 2 shows, this is because ENRICH selected fewer frames and patients from the overrepresented NORMAL class as it chose training subsets that outperformed control sets. When trained on the full training dataset, OCT multi-class performance achieved an AUC of 0.99± 2×10^−4^. Neither ENRICHed nor control subsets were able to achieve this benchmark.

ENRICH outperformed control subsets at just 6 percent of all OCT images (p-value 2.49×10^−9^), with similar findings with representativeness and effective number of classes as in the binary task (**Fig. 2e, Table 2**).

### ECHO-F-SEG multi-class segmentation

The ECHO-F-SEG dataset contains only two images per clip—an intuitive decision to economize on labeling. We evaluated training subsets chosen via ENRICH on multi-class segmentation. Using all available training data (**Table 1**), average Jaccard index was 0.68. With 80 percent of the training data, ENRICH achieved an average Jaccard of 0.66 (**Fig. 2c**). ENRICH did not statistically significantly outperform random selection. Consistent with this, clip-level and patient-level representativeness was statistically indistinguishable between training subsets curated by ENRICH vs. not. (Effective number of classes in this dataset is 1 by definition.)

Model test performance on binary classification in STL10 achieved a mean AUC of 0.99± 2.04×10^−4^ when trained on the full training dataset. In contrast to the medical datasets above, initially, control subsets narrowly but statistically outperformed ENRICH (at 20 percent; p-value 6×10^−4^). At 50 percent of all STL10 images, the trend reversed, and ENRICH narrowly outperformed control subsets (p-value 5×10^−6^) and continued to outperform it as sample size increased. 90 percent of the total dataset was needed in order to achieve gold-standard performance (p-value 0.42; **Fig. 2f**). We were not able to achieve the same benchmark without ENRICH. For multi-class classification with STL10, model test performance achieved a mean AUC of 0.99 (± 5.57×10^−5^) when trained on the full training dataset. Neither subset selection method, ENRICH nor control, was able to achieve the same benchmark. ENRICH was outperformed except at 30 and 40 percent of total, at which sizes performance was indistinguishable. For both binary and multi-class tasks, representativeness and class balance for STL10 were only present at the image level (**Fig. 2f-g**).

### Potential time savings in labeling

W estimated the time required to label all the images in ECHO-F for classification and ECHO-F-SEG for segmentation tasks, from having measured the time it took to manually label these datasets for their original use. We compared this to the time that would have been required for the smallest ENRICHed subsets that achieved desired performance (55 percent for classification and 80 percent for segmentation). Using an ENRICHed training subset would have conferred a savings of 38 hours of full-time work, nearly an entire working week, for an expert labeler, on even this relatively small dataset. Labeling time on OCT and STL10 is unknown.

### ENRICH identifies outliers in image datasets

ENRICH preferentially identifies outlier images, a property that can be exploited as a pre-processing step to screen for noise in large image datasets. In the OCT dataset, for example, we found about 10 percent of images to be similar to each other but different from the rest of images. Investigating this showed that these images had a whitespace padding artifact (**Fig. 3**) that is important to address (e.g. by testing for segregation by class or by using preprocessing or data augmentation) in subsequent model training.

**Figure 3.**
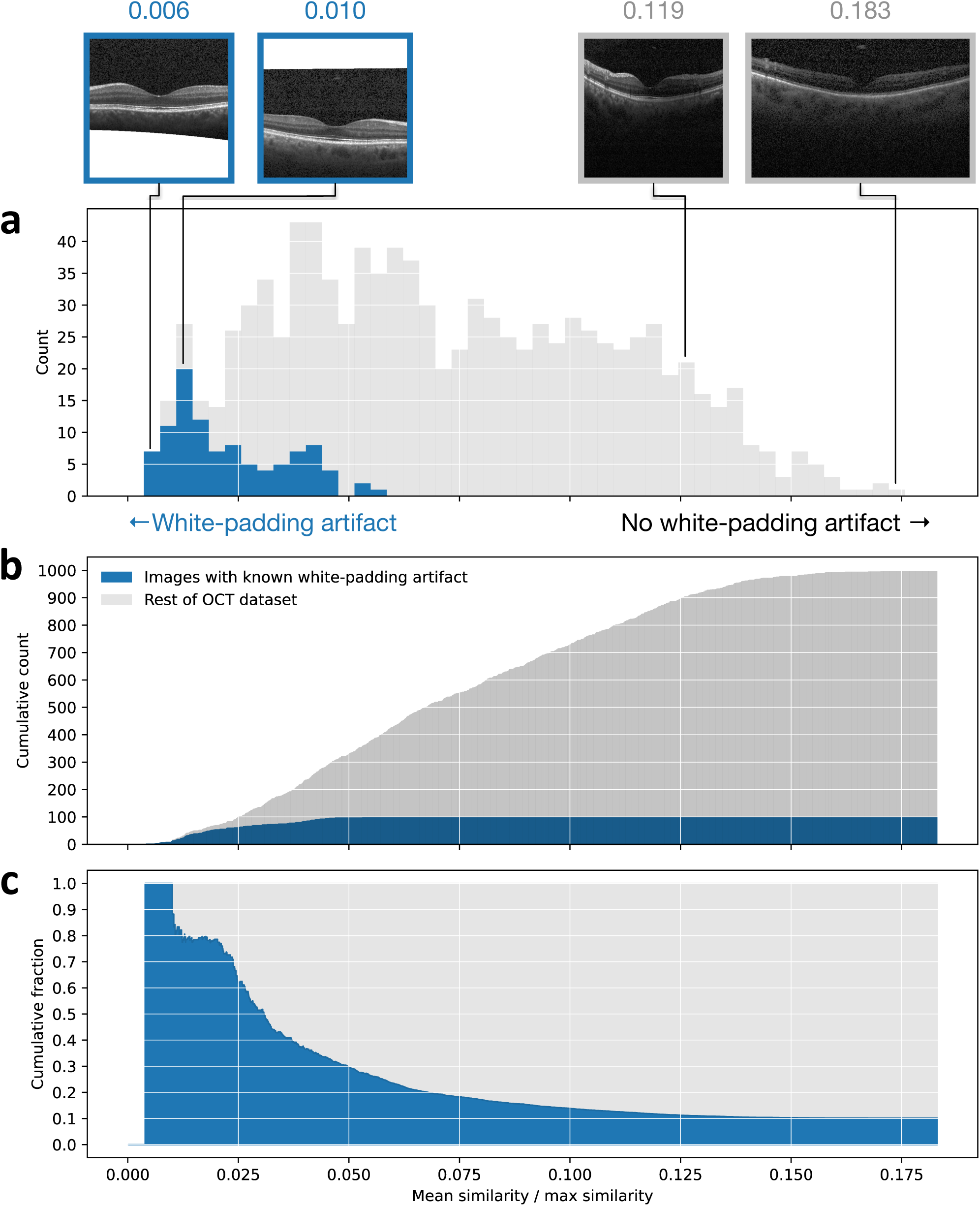
ENRICH aids in screening medical datasets for artifacts. (a) A stacked-bar histogram of mean similarity divided by maximum (non-self) similarity for 1,000 images in OCT. Blue indicates images known to have a white-padding artifact; two examples are shown above, with their mean/max ratio as indicated. (b) Stacked cumulative distribution and (c) cumulative fraction, demonstrating how mean/max facilitates identification of images with artifacts.

Taken together, these data demonstrate that ENRICH can outperform random selection, curating a high-performance training subset that is a fraction of the entire training dataset; and it does so by recognizing structure in the dataset (images, clips, patients) and optimizing both representativeness and class balance for this structure, even though this information is not an explicit part of the current implementation of ENRICH. When such structure does not exist in the dataset—either purposefully removed from medical image data, or in non-medical datasets we encountered—the performance of the current implementation of ENRICH was more modest (**Fig. 2c,f,g**).

## Discussion

In DL for medical imaging, investigators generally rely on a crude metric for dataset quality and content: the number of images in the dataset. With ENRICH, we offer a mathematically rigorous and scalable way to look beyond size to dataset content, a practice that has largely been overlooked or only intuited in DL for medical imaging to date.

Instance selection provides a general strategy for labeling training datasets efficiently. ENRICH curates medical image datasets based on pairwise image similarity. Our results show that ENRICH can be used to identify redundancy in image training datasets. We further demonstrate that medical datasets such as ECHO-F and OCT contain significant redundancy. While the canonical teaching is to split training and test sets by patient, in ECHO-F, most redundancy is at clip level, while in OCT, it appears to be above patient level (perhaps explaining why such a small portion of training data was needed to approximate full training set performance). Additionally, there is a diversity to different classes which mere number of images doesn’t fully describe.

Using ENRICH demonstrated that *(i)* redundant images do not aid significantly in DL model training, *(ii)* this behavior is a property of the image dataset rather than the DL task (binary classification, multi-classification, and segmentation were tested), *(iii)* image labels are not needed in order to curate image datasets according to redundancy nor to optimize representativeness and class balance, *(iv)* images with artifacts can be systematically screened for using pairwise similarities, and *iv)* for some medical datasets, state-of-the-art performance can be achieved using only a fraction of the full training dataset.

For scientific rigor, we demonstrated these findings across several different image datasets, with several replicates per experiment totaling over two thousand model trainings. In some of the experiments presented, performance differences were small in magnitude (but still statistically significant), however, even these can translate to significant dataset savings. Furthermore, performance improvements of a few percentage points often redefine the state of the art when it comes to DL architectures.

There are several points in the above experiments worth mention. First, we note that classification model training demonstrated here did not include standard data augmentations. This choice was made in order to remove data augmentation as a potential confounding factor in measuring the performance of ENRICH. However, in the future, data augmentation can be applied to ENRICH training subsets at the point when they first outperform random selection (e.g. approximately 11 percent of ECHO-F and two percent of OCT), requiring even fewer images to meet optimum test performance.

Second, ENRICH was less helpful for the ECHO-F segmentation task studied here than for the classification tasks. However, for this task, the ECHO-F-SEG dataset had already been intuitively ENRICHed, as only a few image frames per clip were chosen. In this setting, the finding that an additional 20 percent of the already-intuitively-reduced dataset was not needed to reach full dataset performance is still an additional gain in efficiency over informal curation and is another reason that quantitative methods for dataset curation represent an improvement over simple intuition. When considering that labeling each image for segmentation took several minutes, and 20 percent of the training dataset for segmentation comprised 249 image frames, the potential time savings in labeling *even on an already-intuitively-reduced dataset* is significant.

Third, the training/test split for the OCT dataset had to be adjusted in our study because the original test set (500 and 1,000 images in the binary and multi-class datasets respectively) was too easy to classify. Experiments resulted in perfect test set separability (AUC=1.0) despite very small training set sizes (<2 percent total images available). Even with this adjustment, the OCT test dataset was still very separable. In theory, the same methods used in this study to curate training data can be used to curate testing datasets to provide the most efficient and most representative benchmarks for generalizability.

The implications of our findings for economizing on data in DL tasks are clear. Perhaps future studies using medical imaging datasets might benefit from choosing a small, diverse, ENRICHed subset of images to label and use for model development. ENRICH may provide useful metrics on a dataset’s quality and content. While the choice for image similarity metric in the current implementation of ENRICH aided in demonstrating image redundancy as well as noisy images in the dataset, we anticipate that different choices for similarity metric and curation algorithm will yield additional quality metrics. Finally, while many data reduction methods are model-guided: for example in active learning, the model selects the “best” images for learning^36–38^, in an iterative process, ENRICH is data-guided: the data determine which images are best removed, and it can be used once. ENRICH may therefore be used in conjunction with or instead of active learning.

## Conclusion

In the future, investigating alternative similarity measures and ranking algorithms offers opportunities to test and potentially optimize the ENRICH framework. For example, other pairwise image-similarity metrics may prove more informative or simpler to compute. In addition, different algorithm choices as well as code optimizations can be explored to maximize the utility of ENRICH while minimizing time and computational load. Additionally, while we ran over two thousand experiments on several datasets, testing ENRICH on still more datasets can only improve its utility. We make our code available so that others can run ENRICH on their datasets, both for their own benefit and for advancement of the field.

Quantitative measures of similarity have been shown to add useful insights in other fields^35,39^. ENRICH is expected to be a useful new avenue for decreasing labeling burden and speeding iterative training and testing of DL models in development.

## Data Availability

The datasets OCT and STL10 are publicly available at the Mendeley Data repository and the Stanford University Computer Science Department's webpage, https://data.mendeley.com/datasets/rscbjbr9sj/2 and https://cs.stanford.edu/~acoates/stl10/ respectively. Due to patient privacy constraints the ECHO-F and ECHO-F-SEG datasets will not be made available to the public.

## Acknowledgements

EC, RA, RA, and RA were supported by the Department of Defense (W81XWH-19-1-0294) and the National Heart, Lung, and Blood Institute (NIH R01HL146398). RA and RA were supported by the National Institutes of Allergy and Infectious Diseases (NIH R01AI148747-01). RA and RA are supported by the Gordon and Betty Moore Foundation. EC and RA were supported by the American Heart Association (17IGMV33870001).

## Author Contributions

RA and RA conceived of the study. Similarity metric, algorithm design, image preprocessing, and neural-network design and testing were implemented by EC with input from RA and RA. RA, RA, and EC all contributed to the writing of the manuscript.

## Code and Data Availability

Code will be made available at https://github.com/ArnaoutLabUCSF/cardioML upon publication. The datasets OCT and STL10 are publicly available at the Mendeley Data repository and the Stanford University Computer Science Department’s webpage, https://data.mendeley.com/datasets/rscbjbr9sj/2 and https://cs.stanford.edu/~acoates/stl10/ respectively. Due to patient privacy constraints the ECHO-F and ECHO-F-SEG datasets cannot be made available to the public.

